# Effects of Psychedelic Drug Use on Neurocognitive Function and Psychological and Social Quality of Life Domains: An International Online Study

**DOI:** 10.1101/2025.08.27.25334164

**Authors:** Franziska Stadler, Johan Saelens, Ioline Henter, Nathalie Rieser, Maximillian Greenwald, Elizabeth D. Ballard, Katrin H. Preller, Carlos A. Zarate, Christoph Kraus

## Abstract

This international online study (N=759) examined the acute, subacute, and long-term effects of psychedelic drug use on cognitive performance and mental health. Participants completed cognitive tasks assessing working memory, selective attention, and visual/spatial perception, as well as questionnaires assessing mental health outcomes and quality of life. Based on self-reported substance use, participants were classified as non-users, lifetime users, and recent users. Recent users had significantly lower accuracy across all cognitive tasks, and lifetime users had the highest task accuracy without corresponding reaction time deficits. Lifetime use was not associated with long-term cognitive decline. Recent users reported more depressive and dissociative symptoms, whereas lifetime users reported lower scores. Lifetime users scored lower on psychological and social quality of life domains, indicating possible long-term psychosocial effects. These findings highlight the need to differentiate between the acute and long-term effects of psychedelics; lab-controlled, longitudinal studies are needed to enable safe clinical application.

## Introduction

Psychedelic substances such as psilocybin, lysergic acid diethylamide (LSD), mescaline, N,N-dimethyltryptamine (DMT), 2,5-Dimethoxy-4-iodoamphetamine (DOI) and 5-methoxy-DMT (5-MeO-DMT), act primarily by activating the serotonergic 5-HT_2A_ receptor (5-HT2AR) and are referred to as “classic psychedelics” (Rickli et al., 2016; Slocum et al., 2022; Vollenweider et al., 1998), while more dissociative substances such as ketamine and phencyclidine (PCP) primarily act via glutamatergic pathways (Johnston et al., 2023). The past few decades have seen a revived interest in psychedelic-assisted and ketamine-assisted therapy for mental health disorders (Reiff et al., 2020; Vollenweider & Preller, 2020). This renewed focus has led these promising treatments to the verge of regulatory approval (Lamkin, 2022; Mitchell et al., 2021). For instance, esketamine—the *S*-stereoisomer of ketamine—received approval from the United States Food and Drug Administration (FDA) and the European Medicines Agency (EMA) for adults with treatment-resistant depression (TRD) or major depressive disorder (MDD) with suicidal ideation, and 3,4-methylenedioxymethamphetamine (MDMA) was approved by the Australian Therapeutic Goods Administration (TGA) for post-traumatic stress disorder (Haridy, 2023). These clinical developments are supported by population-level findings on psychedelic use, but questions regarding the safety of these psychoactive substances remain. One population study found no significant association between lifetime use of psychedelics and the prevalence of mental health symptoms (including inpatient and outpatient treatment and symptoms of panic disorder, major depressive episode, social phobia, mania and generalized anxiety disorder or psychosis) but did identify a weak positive correlation with better mental health outcomes (T. S. Krebs & P. Ø. Johansen, 2013). Some previous research found no impairment in mental health outcomes following psychedelic drug use (Johnson & Griffiths, 2017; Mans et al., 2021; Studerus et al., 2012). However, to enable more widespread, safe, clinical use, potential adverse events need to be better understood—particularly with regard to cognitive functioning (Bălăeţ, 2022; Bonnieux et al., 2023), as cognition plays a crucial role in psychedelic research due to its overlapping neuronal pathways and its bidirectional relationship with mental health processes.

The 5-HT_2A_ receptor is densely expressed in brain regions associated with cognitive functioning, especially the prefrontal cortex, parietal cortex, and temporal cortex and, to a lesser extent, subcortical regions (Hall et al., 2000; Kadriu et al., 2020; Preller et al., 2019; Saulin et al., 2012; Wong et al., 1987). These areas are integral to memory processing (Bahmani et al., 2019; Williams et al., 2002), executive functioning (Nichols, 2016), visual and spatial perception (Watakabe et al., 2009), and attention (Bahmani et al., 2019), suggesting that psychedelic drug use may have distinct effects on cognition.

Nevertheless, data concerning the acute, subacute, and long-term effects of serotonergic psychedelics on cognitive functioning are scarce. One double-blind, placebo-controlled study exploring the acute effects of a macrodose (100mcg) of LSD on working memory, executive functions and cognitive flexibility, found acute impairment in all domains, that were not seen when taken in combination with the 5-HT_2A_ receptor antagonist ketanserine (Pokorny et al., 2020). Further, two recently published reviews (Basedow et al., 2024; Bonnieux et al., 2023) explored the acute effects of psychedelics, but few data exist on their long-term effects. Likewise, emerging research regarding the cognitive effects of microdosing (broadly defined as ingesting one-tenth of a full dose) has been inconclusive (Kuypers, 2020). One double-blind, placebo-controlled study exploring the effects of microdosing LSD found that it improved sustained attention and did not impair further cognitive functioning, such as working memory, visual processing and cognitive control (Hutten et al., 2019; Hutten et al., 2020). Likewise, other studies investigating the acute effects of microdosing psilocybin or LSD identified no significant impairment or enhancement of cognitive functioning (Cavanna et al., 2022; Family et al., 2019; Szigeti et al., 2021).

Ketamine, another compound with hallucinogenic and dissociative properties, and esketamine (ketamine’s S-stereoisomer) are glutamatergic N-methyl-D-aspartate receptor (NMDAR) antagonists that exhibit a wider expression pattern in the brain (Conti, 1997; Rischka et al., 2022). These receptors are found in brain regions activated during cognitive functioning, suggesting that they may play a role in modulating cognitive processes (Morgan et al., 2010; Zanos et al., 2018), and are also activated in hippocampal neurons mediating synaptic plasticity via long-term potentiation (Vestring et al., 2024). In contrast to the dearth of research regarding the acute effects of serotonergic psychedelics on cognition, several studies have examined the acute effects of ketamine. Short-term ketamine use appears to impair both episodic and working memory (Honey et al., 2005; Morgan et al., 2004), and chronic ketamine users also appear to have impaired cognitive functioning (working memory, visual memory and executive function) (Zhang et al., 2020). One longitudinal study that assessed memory impairment over the course of one year in recreational frequent users (∼20 out of 30 days per month) found significantly impaired memory processing (Morgan et al., 2010). All participants were ketamine users only, as there was a control group with polydrug users. The study also found that ketamine’s effects were dose-dependent, with higher doses of ketamine leading to stronger impairments in cognitive functioning (Zhornitsky et al., 2022). However, ketamine’s impact on cognition might be more complex and results more mixed. In an exploratory, double-blind, placebo-controlled study with 21 individuals with treatment restistant depression, no impairment of working memory, attention and concentration was found after infusion of ketamine (Fijtman, Yavi, et al., 2025). In another randomized, double-blind, placebo-controlled study with 22 participants with treatment resistant depression and 19 healthy participants, the same research team found no effect on working memory after the infusion of one single dose of 0.5mg/kg ketamine (Fijtman, Lally, et al., 2025)

In this context, the impact of classic psychedelics and dissociatives on cognition and mental health calls for further exploration. This international, online study sought to address existing gaps in the data by investigating the potential effects of psychedelics on cognition through cognitive task assessment. Validated questionnaires assessing mental health and quality of life were also administered. The hypothesis was that lifetime or subacute psychedelic drug use would significantly impact working memory, selective attention, and visual perception.

## Results

### Participants and demographic data

Of the 1572 participants who clicked through the study link, 812 participants were excluded for exceeding the time limit of 40 minutes (see Supplement for details regarding the number of excluded participant trials for the cognitive tasks). One participant was excluded because they were underage. Of the 759 participants included in the study, 18.95% (n=144) were in the non-user group, 23.84% (n=181) were in the lifetime user group (meaning they had ingested at least one substance in their lifetime), and 57.2% (n=434) were recent users (defined as participants that ingested at least one substance in the week prior to study participation). From the two user groups, 39.66% (301) participants stated that they were under the influence of a psychedelic drug while completing the study. 7.64% (23) participants of this “currently-using” subsample stated that they had not taken a psychedelic drug in the week prior to the study and were hence counted as lifetime users. To address this confounding factor while still allowing for an analysis with time dependent outcomes, we conducted an additional exploratory analysis excluding the 23 “currently-using” lifetime users, enabling us to better distinguish time dependent drug effects. Participants who stated that they had never used a psychedelic but then listed psychedelic drug use of at least one substance were counted as lifetime users. Demographic data showed that the sample was mostly male (73% male versus 27% female), in early adulthood (32.62 ± 5.93 years), well-educated (80.1% of participants had a tertiary education degree), and from Western countries; 71.5% were US citizens. The most common psychiatric diagnosis among all participants was depression (31.5%), followed by addiction (30.3%) and no mental health condition (26.2%) (Figure 1, Supplemental Table S1).

**Figure 1.**
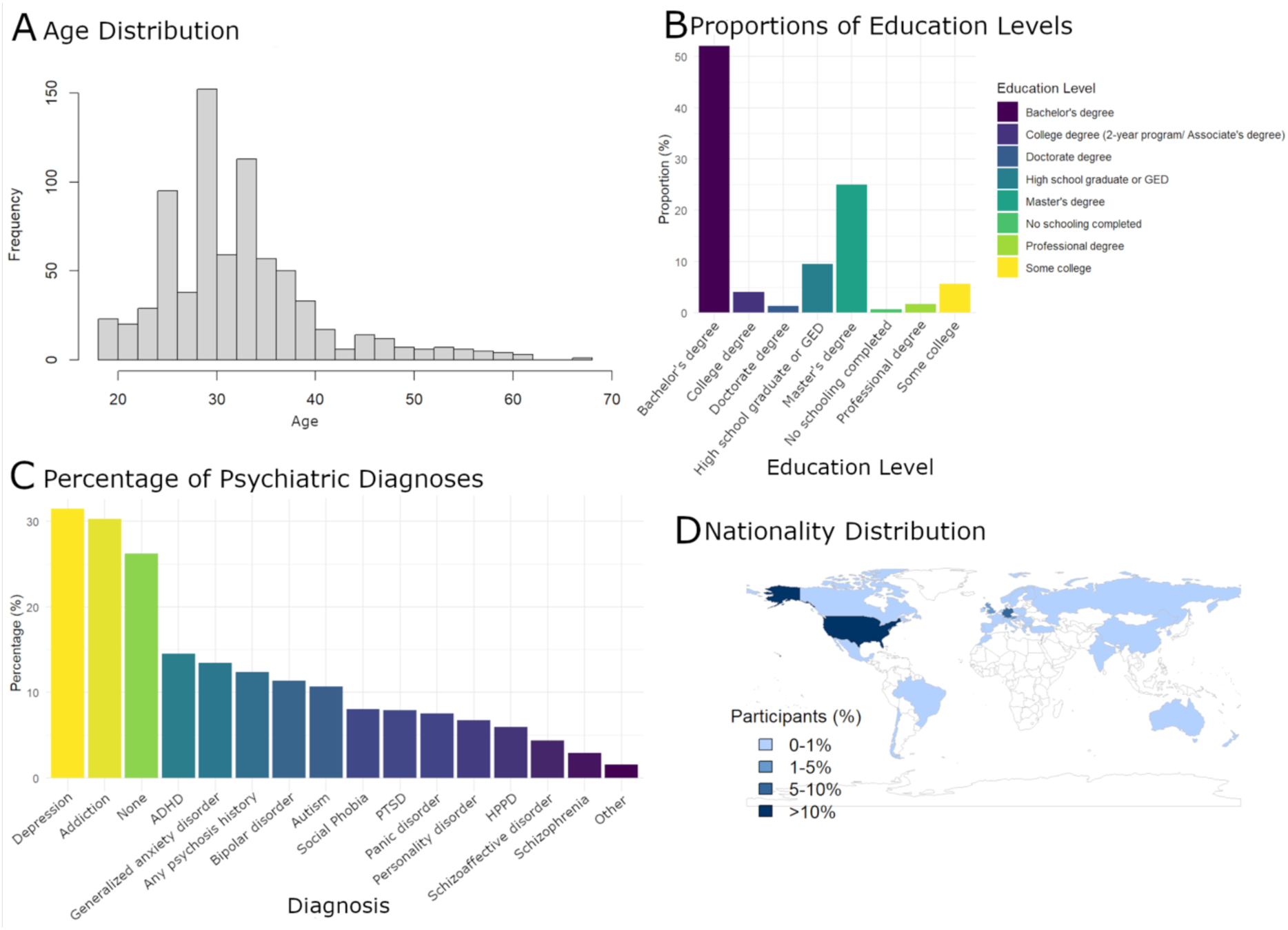
Participant Demographics (n = 759) **A**: The age distribution of participants. **B**: Participants’ education levels. **C**: Percentage of participants reporting psychiatric diagnoses. **D**: World map showing nationality distribution of participants. Darker shades of blue indicate higher percentages of participants originating from a country.

### Drug and alcohol use

With regard to lifetime consumption, the most commonly used drug was alcohol (used by 71.0% of respondents),with a median amount of 130.5 standard drinks per month (a standard drink is approximately 33cl of beer / 15cl of wine / 2cl of spirits (a "shot") / 12 oz of beer / 5oz of wine / 1oz of hard liquor) (interquartile range (IQR) = 407.75 standard drinks). The second most frequently used substance was psilocybin (“magic mushrooms”) (used by 47.0% of respondents); the median dose was 18.0g (IQR = 48.0g) of dried mushrooms, the median number of lifetime uses was 126.5 (IQR = 434.75), and the median amount of times used in the week prior to study participation (recent frequency) was 26.5 (IQR = 51.25). The third most frequently used substance was LSD (used by 46.1% of respondents); the median dose was 125.0mcg (IQR = 166.75mcg), the median number of lifetime uses was 133.0 (IQR = 489.5), and the median recent frequency was 23.0 (IQR = 49.0) (see Figure 2 and supplemental Table S2.) These were followed by, in order of frequency, THC (41.8%, median dose of 25.0g per month (IQR = 27.12g)), cocaine (40.1%, median dose of 34.5g per month (IQR = 26.19g)), MDMA (24.9%, median dose of 100.0mg per month (IQR = 80.0mg), and opioids (23.58%, median dose of 38.0g per month (IQR = 25.95g)).

**Figure 2:**
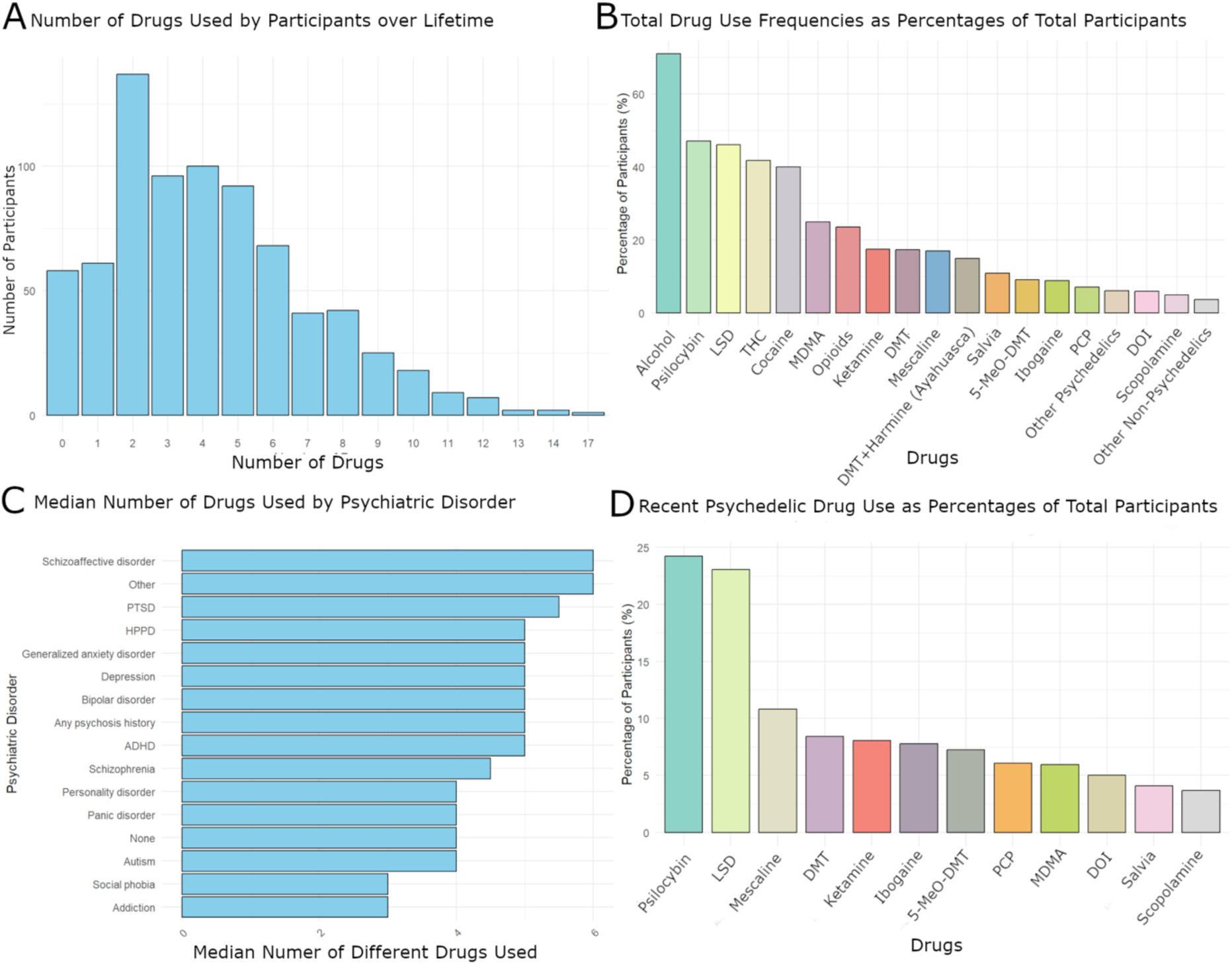
Psychedelics and Additional Drug Use. A: The number of different drugs (psychedelic and other drugs) that participants (n = 759) reported using over their lifetime. B: The percentage of drug use frequencies of participants. C: The median number of different drugs used per participant grouped by psychiatric diagnosis. Participants with schizoaffective disorder and PTSD reported the highest median number of different substances. D: Psychedelic drug use and use of substances with hallucinogenic properties in the week prior to study participation (recent psychedelic drug use).

In this study, 17.5% of respondents reported lifetime ketamine use (median dose of 66.0mg, IQR = 80.0mg, median number of lifetime uses of 240.0 (IQR = 454.5), and median recent frequency of 27.0 (IQR = 30.0). The most common psychedelic drug was psilocybin, followed by LSD and the dissociative ketamine. Other serotonergic psychedelics, such as DMT, mescaline, DMT and harmine (“ayahuasca”), and ibogaine were used less frequently (for dosages and frequencies, see Supplemental Table S2). Most participants reported having used two different substances across their lifetimes, with four and three substances being the next most commonly reported, indicating a prevalent pattern of polydrug use in this sample.

In the week prior to taking part in the study, the most commonly used psychedelic drugs were psilocybin (23.8%) and LSD (22.5%), followed by mescaline (10.6%) and DMT (8.2%), showing a strong preference for serotonergic psychedelics over substances like ketamine (7.5%) or MDMA (5.5%) (see Figure 2).

The most common setting reported for a psychedelic experience was recreational psychedelic drug use (300 participants), followed by psychedelic drug use in the context of research (192 participants) or therapy (173 participants). 216 subjects stated drug experiences with friends. The mean age for first-time psychedelic use was 20.5 years (±8.15 years) (see Supplemental Figure S1).

### Neurocognitive Tasks

Five cognitive tasks were used to measure the three cognitive domains of working memory, selective attention, and spatial/visual perception: the digit span text entry task, the 2-back task, the dot-probe task, the visual search task, and the 2D mental rotation task. The main outcome variables included number of correctly answered trials (accuracy) and the mean reaction time for each cognitive task, meaning that there were 10 variables of interest. Significant differences were observed between the non-user, lifetime user, and recent user groups for most of the 10 main outcome variables (Table 1, Figure 3, for descriptive outcomes see Supplemental Table S3).

**Figure 3.**
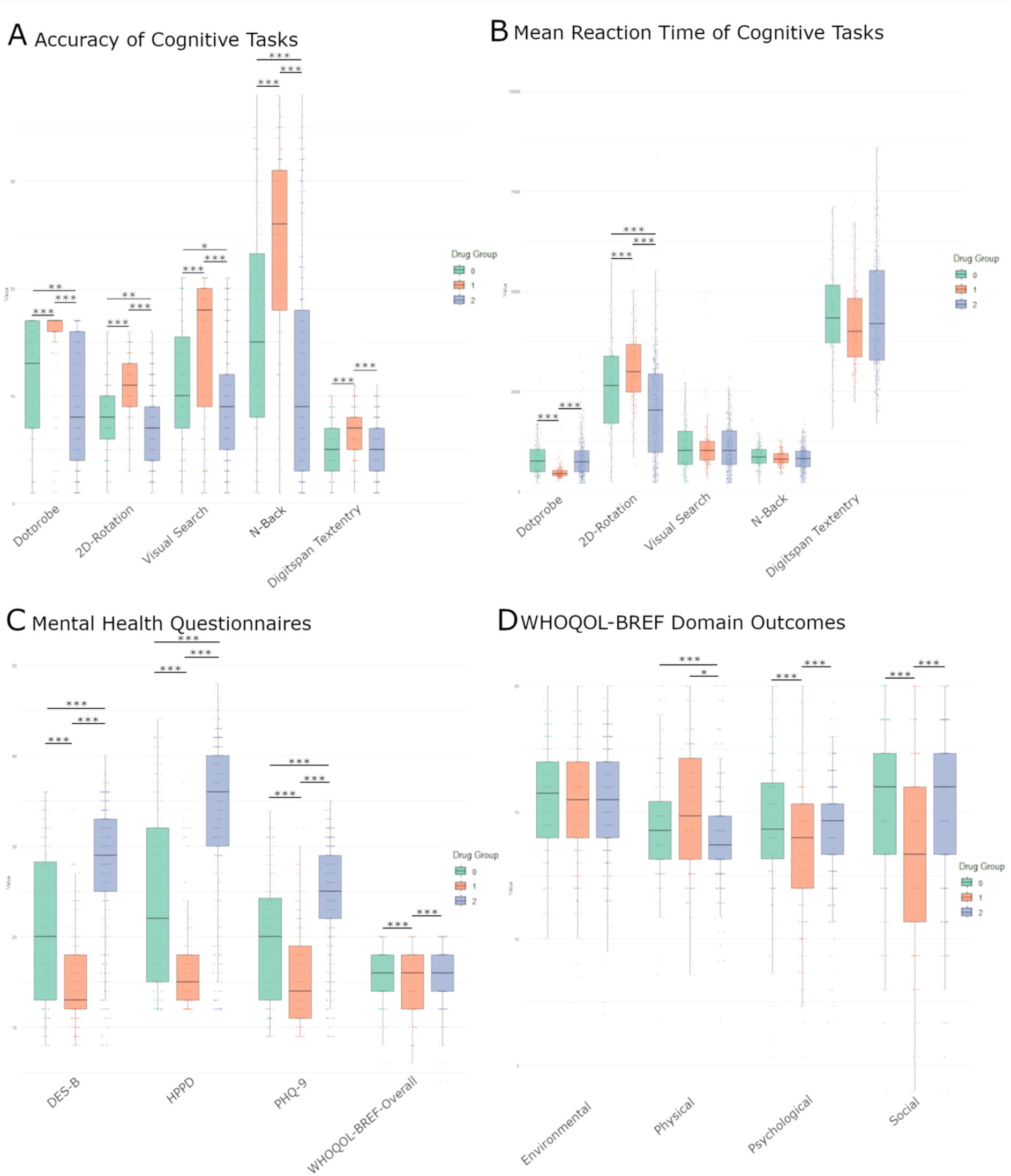
Kruskal-Wallis Test of Cognitive Task Performance and Mental Health by Drug Use Group. Boxplots depicting differences across three drug use groups (n = 759) (0 = non-users (green), 1 = lifetime users (orange), 2 = recent users (blue)) for cognitive performance (accuracy and mean reaction time) and mental health questionnaires. Group differences were assessed using Kruskal-Wallis tests with Dunn’s post-hoc comparisons. Significance is indicated as follows: \**p* < 0.05, \*\**p* < 0.001, \*\*\**p* < 0.0001. **A**: Accuracy of cognitive task performance across five tasks: dot-probe, 2D mental rotation, visual search, n-Back, and digit span text entry. **B**: Mean reaction time for each cognitive task listed in **(A).** **C**: Outcomes from mental health questionnaires, including the Brief Dissociative Experiences Scale (DES-B), Hallucinogen Persisting Perception Disorder scale (HPPD), nine-item Patient Health Questionnaire (PHQ-9), and the overall score from the World Health Organization Quality of Life Questionnaire (WHO-QOL). **D**: WHO-QOL domain-specific outcomes: environmental, physical, psychological, and social.

**Table 1.**
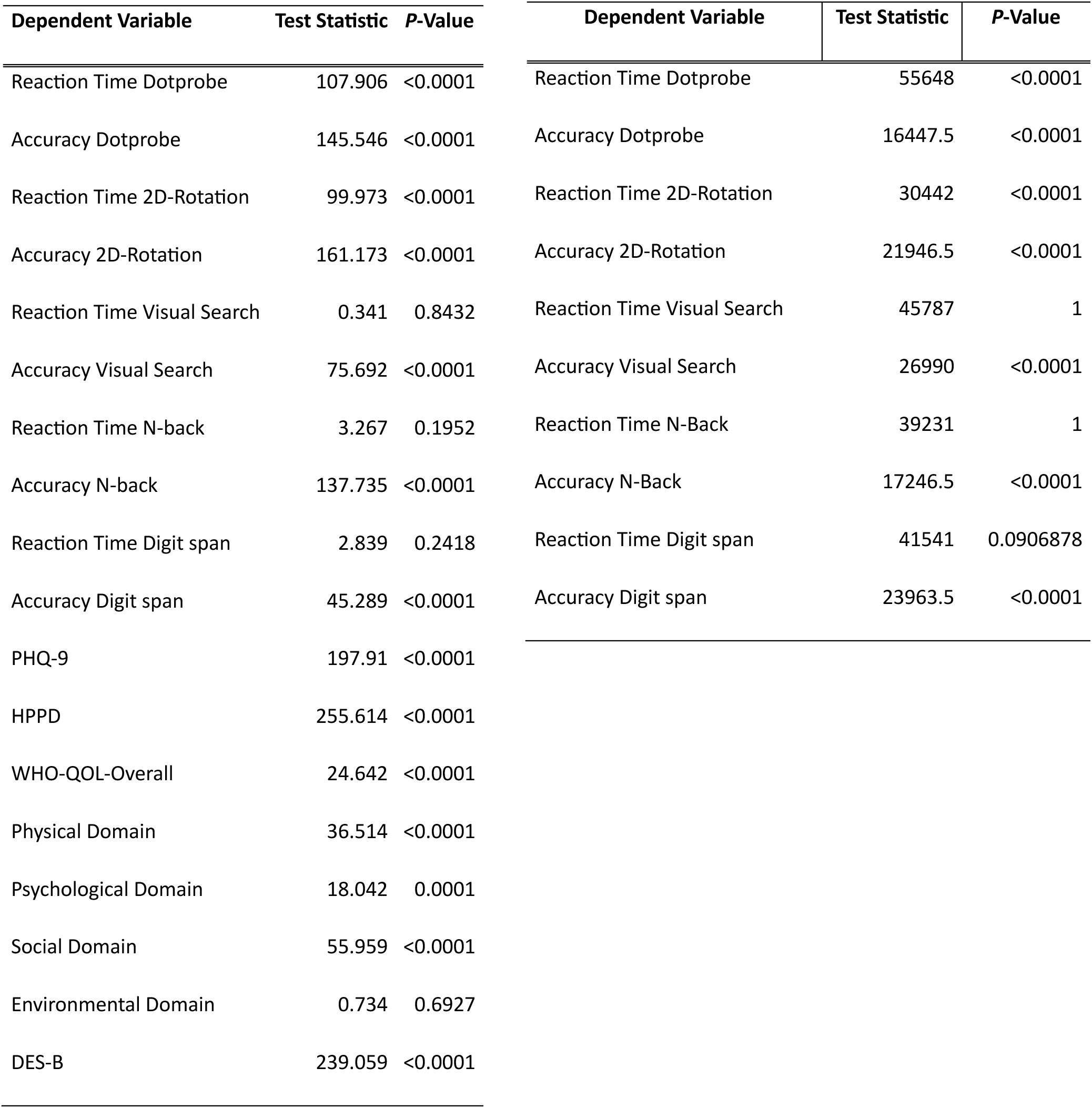
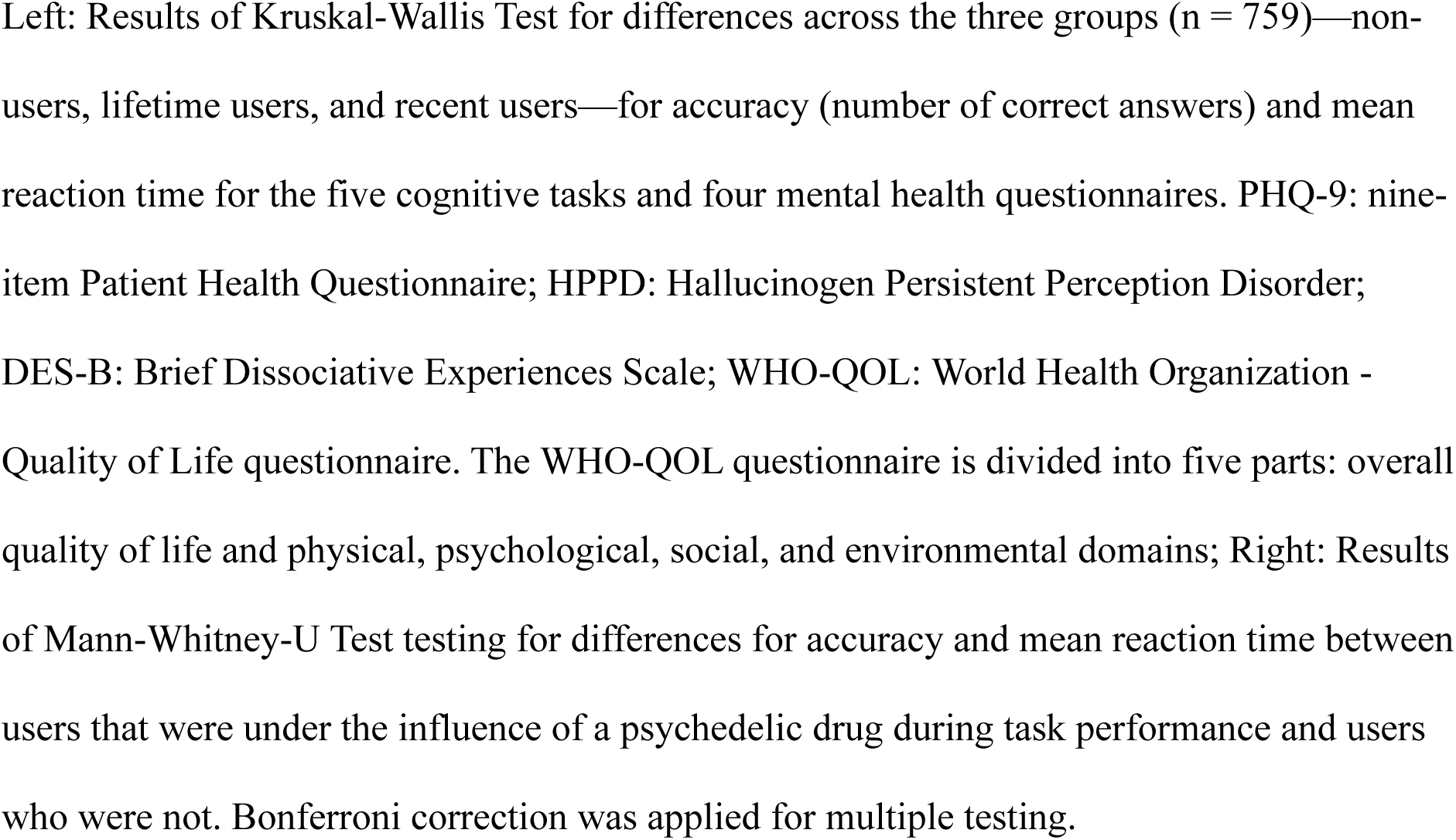
Kruskal-Wallis Task for five cognitive tasks and Mental Health Questionnaires for all participants (left) and Mann-Whitney-U Test for five cognitive tasks between currently-high users and users that were not under the influence of a psychedelic drug during study participation (right)

In the digit span text entry task, which assesses working memory, lifetime users were significantly more accurate than either non-users (*Z* = -4.460, *p* < 0.0001) or recent users (*Z* = 6.627, *p* < 0.0001), while there was no significant difference between non-users and recent users (*Z* = 0.752, *p* = 0.4519). Reaction times did not differ significantly between the three groups (*χ*²(2) = 2.839, *p* = 0.242).

In the 2-back task, which also assesses working memory, reaction times did not significantly differ between the groups (*χ*²(2) = 3.267, *p* = 0.195), but accuracy was significantly higher in lifetime users versus non-users (*Z* = -5.845, *p* < 0.0001) or recent users (*Z* = 11.720, *p* < 0.0001), and recent users performed significantly worse than non-users (*Z* = 3.867, *p* = 0.0001).

Dunn’s post-hoc analysis revealed significant group differences in the dot-probe task, which measures attentional bias, with lifetime users performing significantly more accurately than either non-users (*Z* = -6.694, *p* < 0.0001) or recent users (*Z* = 12.063, *p* < 0.0001). In contrast, recent users performed significantly worse than non-users (*Z* = 3.245, *p* = 0.0012). Lifetime users showed significantly shorter reaction times versus non-users (*Z* = 8.053, *p* < 0.0001) and recent users (*Z* = -9.873, *p* < 0.0001), while no significant difference was found between non-users and recent users (*Z* = 0.276, *p* = 1.000).

In the visual search task, which measures spatial and visual perception, no significant differences were observed in reaction time (*χ*²(2) = 0.341, *p* = 0.843). Lifetime users demonstrated significantly higher accuracy than either non-users (*Z* = 4.418, *p* < 0.0001) or recent users (*Z* = 8.687, *p* < 0.0001). Recent users again performed significantly worse than non-users (*Z* = 2.861, *p* = 0.0042).

Finally, in the 2D mental rotation task—which also evaluates spatial and visual perception—lifetime users had significantly higher accuracy than either non-users (*Z* = - 7.033, *p* < 0.0001) or recent users (*Z* = 12.695, *p* < 0.0001), and recent users showed significantly lower accuracy than non-users (*Z* = 3.517, *p* = 0.0004). With regard to reaction time in the 2D mental rotation task, lifetime users had significantly longer reaction times than non-users (*Z* = -3.770, *p* = 0.0002) or recent users (*Z* = 9.786, *p* < 0.0001), and recent users had significantly shorter reaction times than non-users (*Z* = 4.628, *p* < 0.0001).

Partial correlation testing between task accuracy and recent drug use (defined as consumption of a psychedelic substance within the week prior to study participation) revealed a significant correlation for all five cognitive tasks (dot-probe task: *r* = −0.308, *p* < 0.001; 2D mental rotation task: *r* = −0.331, *p* < 0.001; visual search task: *r* = −0.173, *p* < 0.001; 2-back task: *r* = −0.305, *p* < 0.001; digit span text entry task: *r* = −0.119, *p* = 0.0114). Reaction time outcomes were significantly positively correlated with recent psychedelic drug use for the dot-probe task (r = 0.248, *p* < 0.001) and negatively with the 2D mental rotation task (*r* = −0.240, *p* < 0.001). With regard to the correlation between lifetime use and task accuracy, a significant negative correlation was revealed for the dot-probe task (*r* = -0.155, *p* < 0.001), the 2D mental rotation task (r = -0.166, *p* < 0.001), the visual search task (*r* = -0.119, *p* = 0.011), and the 2-back task (r = -0.223, *p* < 0.001). Mean reaction time was negatively correlated for the 2D mental rotation task (r = -0.187, *p* < 0.001) (Supplemental Table S4 and Figure S2). Corrected Spearman correlations showed dose-dependent effects for LSD, psilocybin, alcohol, and THC for accuracy across all five cognitive tasks, with higher median doses being correlated with lower accuracy. (r= -0.539 to -0.116; p = 0.00694 to p = 9.19e-49). Speed-related correlations showed varying outcomes (Supplemental Table S5). However, a corrected Spearman correlation for drug dose when tested for the covariates of sex, education level, age, and non-hallucinogen use showed no significant association.

To assess the impact of acute psychedelic effects, a Mann-Whitney U-test compared participants who self-reported active psychedelic intoxication during task performance with those who did not. Of 301 participants stating acute psychedelic intoxication, 23 (7.64%) belonged to the lifetime user group and 278 (92.36%) to the recent user group. The first group exhibited significantly lower accuracy across all five cognitive tasks (Table 1 and Figure 4).

**Figure 4.**
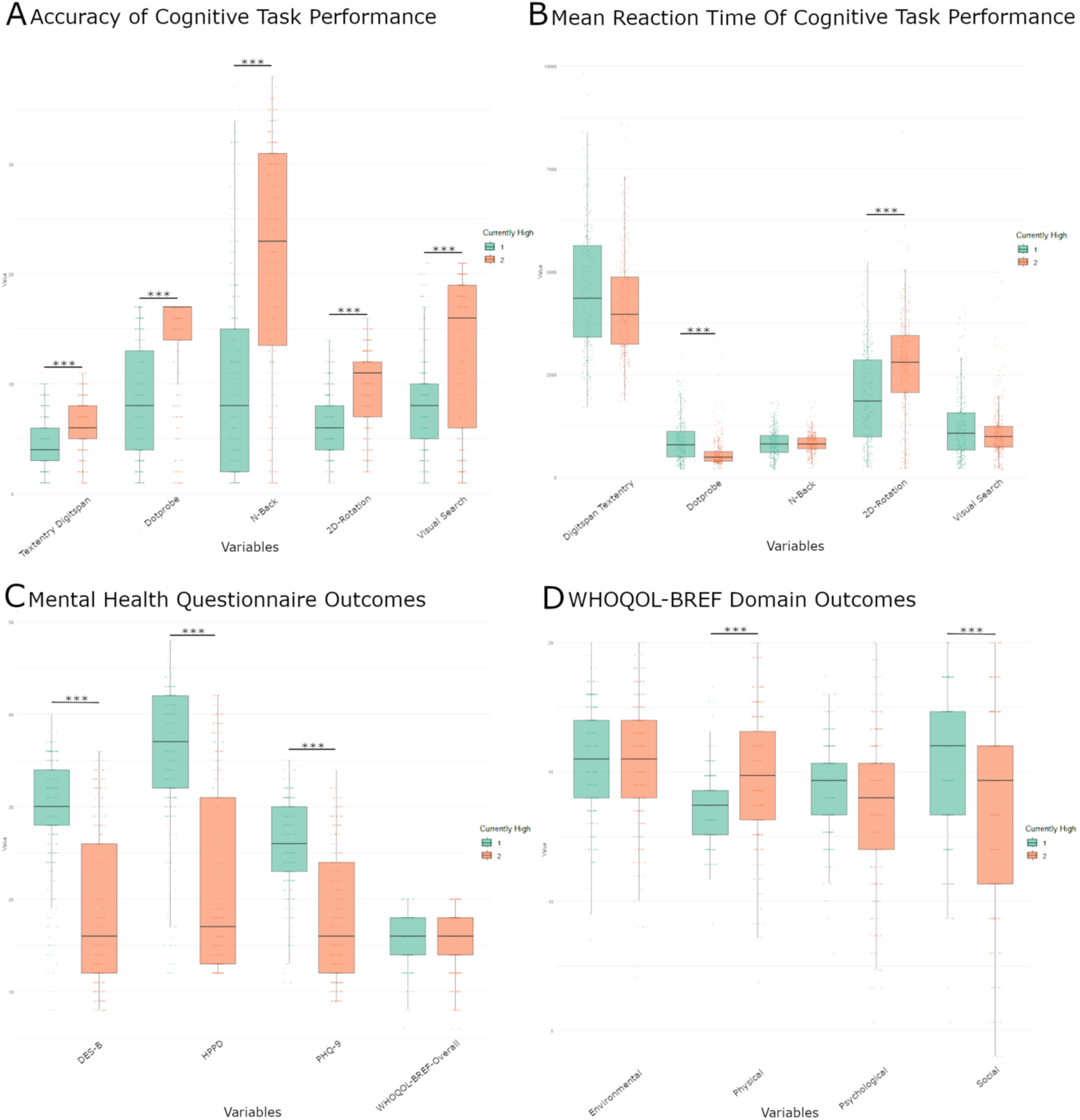
Mann-Whitney U-Test of Cognitive and Mental Health Outcomes grouped by Acute Psychedelic Drug Influence. Boxplots depicting differences between two groups of psychedelic drug users (1=participants who reported that they were under the influence of a psychedelic drug during task performance (green, n = 301), 2=participants who stated that they were NOT under the influence of a psychedelic substance during task performance (orange, n = 314)). Statistical comparison between the groups as performed using the Mann–Whitney U test, with significance levels indicated as follows: *** = *p* < 0.0001. **A:** Accuracy and **B:** mean reaction time of cognitive task performance across five domains: dot-probe, 2D-mental-rotation, visual search, n-Back, and digit span text entry. **C:** Mental health outcomes assessed with the Brief Dissociative Experiences Scale (DES-B), Hallucinogen Persisting Perception Disorder (HPPD) score, and the nine-item Patient Health Questionnaire (PHQ-9). **D:** Self-reported quality of life outcomes measured with the WHO-QOL (World Health Organization-Quality of Life) questionnaire across four domains (environmental, physical, psychological, social) as well as Overall score (**C**).

### Mental health

Group comparisons revealed significant differences in depressive symptoms (Patient Health Questionnaire (PHQ-9); χ²(2) = 197.910, p < 0.0001), Hallucinogen Persistent Perception Disorder (HPPD) scores (χ²(2) = 255.614, p < 0.0001), and dissociative symptoms (Brief Dissociative Experiences Scale (DES-B); χ²(2) = 239.059, p < 0.0001). Across questionnaires, lifetime users showed significantly better mental health outcomes than recent users (Figure 4, for descriptive outcomes see Supplemental Table S6). In addition, post-hoc pairwise comparisons showed that lifetime users had significantly lower depressive symptom scores than both non-users (*Z* = 4.376, *p* < 0.0001) or recent users (*Z* = –13.532, *p* < 0.0001), and that recent users reported significantly higher depression scores than non-users (*Z* = –7.369, *p* < 0.0001). For HPPD, lifetime users scored significantly lower than either non-users (*Z* = 5.563, *p* < 0.001) or recent users (*Z* = -15.537, *p* < 0.0001), with recent users again scoring higher than non-users (*Z* = -7.836, *p* < 0.0001). Similarly, dissociative symptom scores were lowest in lifetime users, who differed significantly from both non-users (*Z* = 5.591, *p* < 0.0001) and recent users (*Z* = -15.077, *p* < 0.0001), while recent users again showed significantly higher dissociative symptoms than non-users (*Z* = -7.380, *p* < 0.0001).

The WHO Quality of Life questionnaire (WHO-QOL) questionnaire is divided into five different domains: physical, psychological, social, environmental, and overall quality of life. Significant differences were found in the physical (*χ*²(2) = 36.514, *p* < 0.0001), social (*χ*²(2) = 55.959, *p* < 0.0001), psychological domains (*χ*²(2) = 18.042, *p* = 0.0001), and overall quality of life (*χ*²(2) = 24.642, *p* < 0.0001), but we found no significant difference in the environmental domain (*χ*²(2) = 0.734, *p* = 0.6927) (Figure 3). Post-hoc pairwise comparisons showed that lifetime users had significantly lower scores in the overall quality of life domain than either non-users (*Z* = 4.381, *p* < 0.0001) or recent users (*Z* = -4.386, *p* < 0.0001), while no difference was observed between non-users and recent users (*Z* = 1.052, *p* = 0.2930). In the psychological and social domains, lifetime users again scored significantly lower than either non-users (psychological: *Z* = 3.615, *p* = 0.0003; social: *Z* = 6.049, *p* < 0.0001) or recent users (psychological: *Z* = -3.870, *p* = 0.0001; social: *Z* = -7.031, *p* < 0.0001), with no significant differences observed between non-users and recent users (psychological: *Z* = 0.637, *p* = 0.5242; social: *Z* = 0.555, *p* = 0.5791). In the physical domain, recent users had significantly lower scores than either non-users (*Z* = 2.761, *p* = 0.0058) or lifetime users (*Z* = 5.922, *p* < 0.0001); the difference between non-users and lifetime users was not significant (*Z* = -2.314, *p* = 0.0207).

Notably, participants who reported being in an acute psychedelic state during task performance showed significantly higher depressive symptom scores (PHQ-9; *U* = 76727.0, *p* < 0.0001), HPPD scores (*U* = 79907.0, *p* < 0.0001), and dissociative symptom scores (DES-B; *U* = 78778.0, *p* < 0.0001) than users who were not acutely intoxicated. They also exhibited higher scores in the physical domain (*U* = 31084.0, *p* < 0.0001) of the WHO-QOL. In contrast, they scored significantly lower in the social domain of the WHO-QOL (*U* = 59415.0, *p* < 0.0001) compared to users who were not in an acute state (Figure 4).

## Discussion

This cross-sectional, international study used online tasks to investigate the acute, subacute, and long-term effects of psychedelic drug use on neurocognitive function and mental health outcomes. By comparing non-users, lifetime users and recent users, the study sought to disentangle the temporally specific impact of psychedelic and additional drug use on cognitive and psychological domains. Our findings provide new insights into the potential cognitive impacts of classic serotonergic psychedelics and ketamine, identifying dose- and time-dependent effects on selective attention, working memory, and visual/spatial perception.

Five cognitive tasks were used to assess the three cognitive domains of interest: working memory, selective attention, and spatial/visual perception. These included the digit span text entry and 2-back tasks for working memory, the dot-probe task for selective attention, and the visual search and 2D mental rotation tasks for spatial/visual perception. Significant differences were observed between non-users, lifetime users, and recent users across all five cognitive task outcomes. Broadly, recent users displayed significantly lower accuracy than non-users and lifetime users, suggesting potential (sub)acute cognitive impairments across the three cognitive domains.

With regard to working memory, these findings support prior research. For instance, one double-blind, placebo-controlled study found that working memory was impaired after high doses (250mcg/kg) of psilocybin (Wittmann et al., 2007), though the sample size was small (n=12) and used a different task (spatial span task) was used to measure working memory. Another double-blind, placebo-controlled study (n=20) also used the 2-back task and found that psilocybin increased reaction time but did not affect accuracy (Barrett et al., 2018). In the present study, lifetime users significantly outperformed non-users and recent users on both tasks that measured working memory (2-back and digit span text entry), though no differences in reaction time were observed. Several limitations and confounding factors listed below prohibit us from drawing the conclusion, that lifetime use might potentially even enhance memory capacity. Additionally, in correlation testing of memory capacity, accuracy was negatively correlated with drug use for lifetime use and recent use.

With regard to selective attention, our findings of (sub)acute impairment in recent users and increased accuracy in lifetime users are in line with previous research, though data on attentional bias are scarce (Basedow et al., 2024). Most notably, 39.66% (n=301) of the participants, all of whom belonged to the lifetime or recent user groups, stated that they were under the influence of a psychedelic drug while taking part in the study, preventing us from differentiating between acute and subacute effects. Impairment might thus appear as an acute effect, given that prior studies observed no subacute impairments in attention. For example, one open-label study of 24 participants with major depressive disorder found no impairment in selective attention one and four weeks after administration of one moderate-dose (20mg/kg) and one high-dose (30mg/kg) of psilocybin (Doss et al., 2021). Another double-blind, placebo-controlled study found no subacute impairment of attention 24 hours after LSD administration (50mcg) (Wießner et al., 2022). However, most research has examined attentional bias in the context of microdosing and obtained mixed results ranging from reports of no observable effects (Family et al., 2019) to enhanced attentional performance (Hutten et al., 2020). These inconsistencies highlight the methodological challenges associated with comparing studies that lack standardization, particularly concerning varying doses, sample sizes, and cognitive tasks (Bălăeţ, 2022). Further research investigating other domains of cognition show equally varying outcomes. For example, in the aforementioned study by Doss and colleagues (2021), improved cognitive flexibility was observed after two macrodose psilocybin (20mg/kg and 30mg/kg, respectively) administrations, whereas a double-blind, placebo-controlled study by Mason and colleagues noted acutely impaired creative thinking after administration of a macrodose of psilocybin (0.17mg/kg), followed by increased creative thinking seven days later (Mason et al., 2021).

When considering dose-dependent effects, no significant associations were noted between dose and cognitive task outcomes after controlling for covariates. However, analyses focusing on individual substances—LSD, psilocybin, alcohol, and THC—consistently found that higher doses were associated with impairments across all three cognitive domains. This is in line with previous research suggesting that high doses impair cognitive functioning, while lower doses may facilitate enhancement (Basedow et al., 2024; Bonnieux et al., 2023). Nevertheless, longitudinal and more consistent study designs are needed before conclusions can be drawn beyond acute effects only (Bălăeţ, 2022).

Interestingly, lifetime users demonstrated higher accuracy than non-users or recent users across all five cognitive tasks. However, these findings were not supported during correlation testing. Thus, the findings do not suggest a potential long-term cognitive benefit associated with psychedelic experiences, paralleling previous research that found that psychedelics had no long-term effects on cognition (Bonnieux et al., 2023; Rucker et al., 2022). In addition, because participants who reported being under the influence of a psychedelic drug during the cognitive task were included in the lifetime users group, it is difficult to disentangle acute from long-term effects. To address this bias, we performed the statistical analysis after excluding the 23 “currently-using” lifetime user group subjects in an explorative approach (see Supplement Table S7 and S8). The outcomes of the Kruskal-Wallis test and the post hoc Dunn’s test remained unchanged, as did the results of the correlation analyses. This allows us to interpret the findings as previously described, considering lifetime users as such without the confounding factor of current intoxication.

Interestingly, recent users in this analysis had significantly higher depressive symptom scores (as assessed by the PHQ-9), and a higher prevalence of HPPD symptoms and dissociative symptoms (as assessed by the DES-B) than either non-users or lifetime users, possibly reflecting that acute or subacute aftereffects might cause psychological distress. It is also important to take the bilateral relationship between mental health and psychedelic substance use into account, as participants with mental health conditions might also have turned to psychedelic experiences in an attempt of self-medication or in a therapeutic or research context. It should also be noted that the prevalence of depressive disorders in our sample was much higher (31.5%) than typically observed in a population study; for instance, one US study reported a prevalence of 18.4% in 2020 (Lee et al., 2023). Notably, lifetime users in this study had significantly lower depressive symptom scores, fewer dissociative symptoms, and lower HPPD scores than either recent users or non-users. While previous research found no negative long-term mental health outcomes in psychedelic drug users (Johnson et al., 2019; T. S. Krebs & P. Ø. Johansen, 2013), it is not possible to draw the same consclusion, as recent users who showed higher scores might also be long-term users.

It should also be noted that in the WHO-QOL, lifetime users scored lower in the psychological and social domains than non-users or recent users, potentially suggesting that psychedelic use has long-term social effects. These outcomes might be due to difficulties integrating challenging psychedelic experiences or altered social behavior. However, these findings may also be influenced by selection bias, as individuals with a strong interest in psychedelics or particularly positive or negative experiences may have been more likely to participate in the study.

Despite the strengths of our study—which include its international, naturalistic design and easy accessibility—several limitations bear mention. First, in an online study we have little control over the setting participants undergo during questionnaires and tasks and considerable variability may exist with regard to the type of devices used or other environmental distractions, leading to variability in the dataset. Second, potential biases might be present when self-reporting drug use and mental health symptoms, either as a result of difficulties recalling substance or dose accuracy or because of potential social pressure. As a salient example, the psilocybin content of specific species of psilocybin-containing mushrooms often remain unknown to users; psilocybe cubensis, the most commonly used species, contains approximately 10 mg of psilocybin per gram of dried mushroom (Clarke, 2007), which served as the reference dose in this study. Third, while this study assessed additional drug use, it did not take into account nicotine use or prescribed medications, including those with neurocognitive (side) effects. Fourth, some recent, as well as lifetime users reported being under the influence during study participation, which limited our ability to differentiate between acute and subacute effects. We addressed this bias by conducting an exploratory additional analysis, excluding the “currently-using” participants from the lifetime user group. While this enabled us to differentiate between long-term and (sub)acute effects, differentiation between acute and subacute effects remained limited. Furthermore, the arbitrary nature of our group configuration limits application on real life scenarios, as participants who stated psychedelic drug use for example seven days before study participation were counted as recent users, but participants who stated psychedelic drug use eight days prior were counted as life-time users. Also, the total amount of substances used is not reflected in the outcomes of the Kruskal-Wallis test, not taking into account that recent users are most likely the subgroup with the heaviest drug use history. Fifth, like most psychedelic research, a selection bias may exist regarding the prevalence of psychedelic drug use in association with socioeconomic and sex differences. Population-based studies suggest that the prevalence of lifetime psychedelic drug use is 17% between the ages of 21 and 64 (T. S. Krebs & P.-Ø. Johansen, 2013), with 16.1% reporting psilocybin use in the past 12 months (Winstock Ar et al., 2021), revealing much lower prevalence rates than in our study (47% of participants reporting lifetime psilocybin use and 46.1% of participants LSD lifetimes use, respectively. This difference might stem from our recruitment strategies, which specifically targeted participants interested in psychedelic research and who were thus potentially more open to psychedelic experiences. Recruitment strategies might have also skewed data concerning age, as especially the platform “Reddit” is more likely used by younger adults. Sixth, there is a potential selection bias in that chronic psychedelic users are likely those who have not experienced adverse effects, as individuals with negative initial experiences are less likely to continue use, potentially limiting the representativeness of frequent users for the average psychedelic experience. Our sample was homogenous, well educated, primarily male, and from predominantly western countries. While this might not be an accurate reflection of the average psychedelic drug user, most research into psychedelics have observed similar participant samples (Bonnieux et al., 2023), suggesting that more diverse participant samples and adjusted recruitment strategies will be crucial for establishing a safety profile for these agents across the general population. To address some of these limitations, our analyses included covariates such as psychiatric conditions, age at first use, age, sex, use of non-hallucinogens, and education level and used validated cognitive tasks.

Despite these limitations, this large-scale, naturalistic study offers novel insights into the cognitive and psychological correlates of psychedelic use by distinguishing between non-users, recent users, and lifetime users across multiple cognitive domains. Our findings provide evidence of (sub)acute cognitive impairments in the group of recent users in the three cognitive domains of working memory, selective attention, and visual/spatial perception; specifically, recent use was associated with reduced task accuracy. Lifetime users, in contrast, demonstrated superior performance in several cognitive tasks compared to the other groups. However, these apparent benefits were not supported by correlation analyses testing for correlation between the total amount of psychedelics and dissociatives and cognitive task outcomes, suggesting that lifetime use does not enhance long-term cognitive advantages. Furthermore, while recent users reported elevated levels of depressive, dissociation, and HPPD symptoms, lifetime users reported fewer symptoms than either of the other groups, aligning with previous findings that psychedelics have no long-term adverse mental health effects. Nevertheless, lower scores on psychological and social quality-of-life domains among lifetime users raise the possibility of subtle, long-term psychosocial consequences, potentially related to the integration of challenging psychedelic experiences. Alternatively, the observed long-term effects may reflect pre-existing psychological vulnerabilities in individuals who turned to psychedelics for help, rather than consequences of psychedelic use itself.

In conclusion, this international online study examined the acute, subacute, and long-term effects of psychedelic drug use on cognitive performance and mental health. Its strengths included a large, international sample and easy accessibility. Nevertheless, this study was limited by its cross-sectional design, reliance on self-reported drug use and mental health symptoms, and potential confounding from acute intoxication during participation. The overrepresentation of highly educated, western, and male participants also limits generalizability. Nevertheless, the study found that recent users had significantly lower accuracy across all cognitive tasks, and lifetime users had the highest task accuracy without corresponding reaction time deficits. Lifetime use was not associated with long-term cognitive decline. Recent users also reported more depressive and dissociative symptoms, whereas lifetime users reported lower scores. Future longitudinal, lab-, and placebo-controlled designs with standardized dosing and diverse, representative samples should be used to deepen our understanding of the complex and dose-dependent cognitive and psychological consequences of psychedelic use.

## Materials and Methods

### Study design

This is a naturalistic, international online study evaluating the effects of psychedelic drug use on cognitive functioning. It was conducted on the platform Gorilla (www.gorilla.sc), an online experiment builder that allows reliable online measurement of neurocognitive tasks (Anwyl-Irvine et al., 2021; Anwyl-Irvine et al., 2020). For feasibility reasons, a set of questionnaires and tasks that could be filled out during an approximately 30-minute period was used. Ethical approval was obtained by the Ethical Commission of the Medical University of Vienna, Austria (reference number: 1618/2021), and the study was registered on www.osf.io (reference: https://doi.org/10.17605/OSF.IO/N7UHX). Participants gave consent of study design and data usage by agreeing to a consent form at the beginning of the study after the landing page.

### Participants and Recruitment

The study recruited participants worldwide who were over the age of 18 and who accessed the study via a laptop or PC; the study specifically targeted people interested in the realm of psychedelics. We included all questionnaires that were complete, carefully filled out and in the allotted amount of time. There were no specific exclusion criteria apart from minimum age of participation and outcomes with implausible answers or reaction times.

Participants were recruited between February 2022 and December 2024. A landing page was generated where participants could access the study and, if interested, were redirected to gorilla.sc. Recruitment strategies included advertisement on social media platforms (Instagram, Facebook and Twitter) as well as promotion on the Reddit platform, where we actively sought participants interested in psychedelics in subreddits dedicated to specific topics, such as “psychonaut”, “mushrooms”, “HPPD”, “RationalPsychonaut”, “microdosing”, “psychedelics”, “LSD”, “depressionregimens” and “PsychedelicTherapy”. The study was also promoted via the third party provider Amazon Mechanical Turk, a platform that links researchers and participants; participants who took part via Amazon Mechanical Turk were financially reimbursed with $5 USD.

### Procedure

Participants were first asked for their demographic data as well as mental health conditions, indicating any psychiatric diagnoses, their history of inpatient psychiatric care, history of drug or alcohol use, and hospitalization for psychoactive drug-related incidents.

Participants were then questioned regarding their history of psychedelic drug use. Options included serotonergic psychedelics (psilocybin, LSD, DMT, tryptamine (5-MeO-DMT), mescaline, ibogaine, (2,5-Dimethoxy-4-iodoamphetamine (DOI), and substances with psychedelic properties, such as the glutamatergic dissociatives ketamine and phencyclidine (PCP), and substances exerting their effects via different pharmacological mechanisms, such as scopolamine, salvia, and MDMA (Kalant, 2001). Data on further drug use, such as alcohol, cocaine, cannabinoids, and opiates were also collected. Participants reported the frequency of use within the past month, the frequency of use during their period of highest use, the number of classic psychedelics and substances with hallucinogenic properties used both in their lifetime and the past week, and the median dose of each used substance. Age of first use of a psychedelic was also assessed.

### Cognitive Tasks

Five cognitive tasks were used to assess the three cognitive domains: working memory, selective attention, and spatial/visual perception. The n-back task (in this study a 2-back task), and the digit span text entry task were used to assess working memory. Selective attention was measured using the dot-probe task, which captures attentional shifts in response to visual stimuli (Koster et al., 2004). For visual and spatial perception, the 2D mental rotation task (Vandenberg & Kuse, 1978) and the visual search task were used. A more detailed description of the task procedures is available in the Supplement. All tasks were provided by the Gorilla platform (www.gorilla.sc), although trial answers were partly designed by the research team (see Supplement). Task reaction times and number of correct answers were recorded using Gorilla.sc, ensuring timing accuracy comparable to lab-based setups through high-resolution timers and frame counting (Anwyl-Irvine et al., 2021). Responses were given manually on the keyboards of the participant’s devices. Before each task, participants completed a test trial.

Participant data with reaction time less than 200ms or above 10000ms of reaction time were removed only for the task in question (the dot-probe task and 2-back task had implemented maximum time limits). Only the trial in question was excluded, while the rest of the participant’s data was included in analysis.

### Questionnaires

Participants completed four questionnaires concerning mental health and quality of life. Symptoms of depression were assessed using the PHQ-9 (Martin et al., 2006), overall quality of life and health was evaluated with the WHO-QOL (Harper et al., 1998), and dissociative symptoms were measured by the DES-B (Carlson et al., 2018). To assess clinical symptoms of HPPD, a questionnaire (Locke) designed by Steven Locke, MD was used that has 12 items measuring symptoms experienced within the month prior to participation; these focus on distortions of visual perception such as colors, geometric patterns, shapes, and light. Participants responded to each question using a scale with options ranging from “not at all”, “several days”, and “more than half the days” to “nearly every day”.

### Statistical analysis

Data were extracted from Gorilla (www.gorilla.sc) via spreadsheets and analyzed in Rstudio (version 4.4.1) (Team, 2021). One participant who did not meet inclusion criteria because they were underage was removed from the dataset. Extreme outliers in the cognitive tasks—those with a reaction time above 10s or lower than 200ms—were also removed for each trial in question (see Supplement).

Demographic data and mental health diagnoses were analyzed using descriptive statistics including mean, median, standard deviation, minimum, and maximum of the metric variables. Prior to data collection, we had planned to divide participants into those using serotonergic psychedelics, glutamatergic agents, or other agents, with the intention of comparing them to a control group of non-users. This stratification sought to differentiate the cognitive effects of serotonergic versus glutamatergic agents, given the distinct expression patterns of their main target receptors in the brain. However, during data collection, it became evident that the number of participants who exclusively used glutamatergic substances such as ketamine or PCP was too small to allow statistical analysis. This may have been due to the strong dissociative effects of ketamine, which make it less likely to serve as an initial substance of use or because our sample predominantly comprised US citizens, where ketamine use is less prevalent (SAMHSA, 2022). Consequently, we decided post-hoc to divide the participants into three groups: non-users (defined as participants who stated they had never previously used a psychedelic drug); lifetime users (defined as those who had used at least one psychedelic drug at least once in their lives); and recent users (defined as participants who had taken at least one psychedelic drug in the week prior to taking part in the study. Furthermore, some participants across both groups of users stated that they were under the influence of a psychedelic drug while completing the study tasks (hereafter referred to as “currently-using” users); this group of “currently-using” users was compared to all users not under the influence of a psychedelic drug regardless their previous psychedelic drug use history, to test for acute effects associated with psychedelic drug use. As 23 participants who reported lifetime use and acute psychedelic intoxication during study participation also indicated no psychedelic substance use within the week preceding participation and were therefore classified as lifetime users, we conducted an exploratory re-analysis excluding these 23 participants (see Supplement Table S7 and S8).

All statistical tests were performed to differentiate between the acute, subacute, and long-term effects of psychedelics on task performance and mental health outcomes. To evaluate differences between the main outcome variables (accuracy and reaction time for each cognitive task), the Kruskal-Wallis test was employed, as data were skewed. Post-hoc Dunn’s tests were applied for significantly different variables between the three groups. Because data were not normally distributed, a correlation between cognitive outcomes and drug use was calculated with partial Spearman’s correlations, with age at first use, sex, education level, age, use of non-psychedelics, and mental health diagnoses as covariates. Spearman correlations were further used to test for correlations between cognitive outcomes and average drug dose, with sex, education level, age, and use of non-psychedelics as covariates. Mann-Whitney-U-tests were used to test for differences in cognitive outcomes and mental health outcomes between the group configuration of “currently-using” users and users not under the influence of a psychedelic drug during the tasks. To test for differences in the mental health questionnaires, the Kruskal-Wallis test followed by Dunn’s tests were applied. Bonferroni correction was used to correct for multiple testing for Post-hoc Dunn’s test and for Mann-Whitney-U-tests. Dose-dependent task outcomes were tested via Spearman correlations for the most used drugs (LSD, psilocybin, alcohol, and THC).

## Data Availability

All data produced in the present study are available upon reasonable request to the authors
All data produced in the present work are contained in the manuscript
All data produced are available online at osf.io (https://doi.org/10.17605/OSF.IO/N7UHX)

## Acknowledgements

Ioline Henter (NIMH) provided invaluable editorial assistance.

