## supplemental tables and figures for "Effects of Psychedelic Drug Use on Neurocognitive Function and Psychological and Social Quality of Life Domains: An International Online Study"

### Supplement

Demographic data asked included age, sex, nationality, highest completed education level, years of education, employment status, work hours per week, number of jobs, annual income in US dollars, kind of residence, marital status, number of children currently being cared for, and level of disability.

Five cognitive tasks provided by Gorilla.sc were used to measure working memory, selective attention, and spatial/visual perception.

**N-back task:** In the n-back task, which measures working memory, participants must recall the letter presented n trials earlier. In this version of the task, a 2-back task, participants had to remember the letter shown two trials prior. If no response was given within 2000 ms, the task automatically moved to the next trial, and the trial was considered incorrect. Performance was measured based on the number of correct answers and reaction time. The task consisted of 35 total trials.

**Digit span text entry task:** In the digit span text entry task, which measures working memory, participants were shown a series of digits they had to remember in the correct order. They were then asked to type in the remembered digits. The task included 10 trials, with each trial featuring a maximum of eight digits. Performance was measured based on the number of correct trials and reaction time.

**Dot-probe task**: The dot-probe task measures selective attention (Koster et al., 2004). Two pictures were presented simultaneously, and a dot appeared behind one of them. Participants had to identify the picture containing the dot as quickly as possible. The pictures were shown for 200 ms, and if no response was given within 5000 ms, the task automatically moved to the next trial, and the trial was considered incorrect. Performance was evaluated based on accuracy and reaction time. The task included 16 trials.

**2D mental rotation task:** We measured visual and spatial perception using the 2D mental rotation task (Vandenberg & Kuse, 1978). Participants had to identify the two-dimensionally rotated version of a target object. The target object was displayed for a maximum of 1500 ms. Performance was measured by number of correct trials and reaction time. The task consisted of 16 trials. Task performance is associated with activation of the parietal lobe (Gogos et al., 2010).

**Visual search task:** In the visual search task, which measures visual and spatial perception, participants had to locate an item characterized by two specific features (color and shape) within a group of items, each displaying only one of the two features. The items were presented for 600 ms. Performance was measured based on the number of correct trials and reaction time. The task included 20 trials. Brain regions involved in visual search include the primary visual cortex, the right posterior parietal cortex, the right dorsolateral prefrontal cortex, and the dorsal anterior cingulate cortex (Bueichekú et al., 2019).

For each task, trial answers were partly designed by the platform itself and partly by the research team. Specifically, in the 2D mental rotation task, three trials were designed by www.gorilla.sc, while 13 were designed by the research team. For the visual search task, 12 trial stimuli were provided by www.gorilla.sc, while the researchers designed eight. In the dot-probe task, gorilla.sc provided stimuli for eight trials, while the research designed eight remaining trials. Finally, in the 2-back task and the digit span text entry task the research team designed all the letter sequences as well as the digit sequences.

Participant data were excluded for each trial when reaction times were lower than 200ms or higher than 10,000ms. In such cases, only data from the relevant trial were excluded, not all the participant's data. In addition, the dot-probe and 2-back tasks had implemented maximum time limits, leading to exclusion only because of too quick reaction times. For the dot-probe task, 2449 (20.05%) of the trials were excluded; for the n-back task, 6119 (18.3%) of the trials were excluded; for the 2D mental rotation task, 1709 (10.2%) of the trials were excluded (458 (2.7%) for responding too quickly and 1252 (7.5%) for responding too slowly); for the visual search task, 4578 (12.8%) of the trials were excluded (3126 (8.7%) for reacting too quickly and 1452 (4.1%) for reacting too slowly); finally, for the digit span text entry task, 18784 (17.7%) of the participant trials were excluded (16598 (15.6%) for responding too quickly and 2186 (2.1%) for responding too slowly).

During the analysis of average drug dose and frequencies, participants who stated "0", meaning that they had never taken that substance, for each respective substance, were excluded in order to get less skewed data.

**Supplemental Table S1. Demographic and Clinical Characteristics**

| Demographics | Total Participants N=759 |
|---|---|
| **Age** | |
| Median | 32 |
| Mean | 32.62 |
| Min-Max | 18-67 |
| Missing Age (NA) | 2 |

| | |
|---|---|
| **Sex** | |
| Female – no. (%) | 205 (27.01) |
| Male – no.(%) | 554 (72.99) |
| **Nationality** | |
| US American – no. (%) | 543 (71.5) |
| **Education** | |
| Tertiary Education Degree – no. (%) | 608 (80.1) |
| **Psychiatric Condition** | |
| Depression – no. (%) | 239 (31.49) |
| Addiction – no. (%) | 230 (30.30) |
| None – no. (%) | 199 (26.22) |
| ADHD – no. (%) | 110 (14.49) |
| Generalized anxiety disorder – no. (%) | 102 (13.44) |
| Any psychosis history – no. (%) | 94 (12.38) |
| Bipolar disorder – no. (%) | 86 (11.33) |
| Autism – no. (%) | 81 (10.67) |
| Social phobia – no. (%) | 61 (8.04) |
| PTSD – no. (%) | 60 (7.91) |
| Panic disorder – no. (%) | 57 (7.51) |
| Personality disorder – no. (%) | 51 (6.72) |

| | |
|---|---|
| HPPD – no. (%) | 45 (5.93) |
| Schizoaffective disorder – no. (%) | 33 (4.35) |
| Schizophrenia – no. (%) | 22 (2.90) |
| Other – no. (%) | 12 (1.58) |

Age in years; No.: Number;

### Supplemental Table S2: Assessment of classic psychedelic drug use and use of substances with hallucinogenic properties

| Drug | Median Dose | Mean Dose | SD Dose | IQR Dose | Median Lifetime Use | Mean Lifetime Use | SD Lifetime Use | IQR Lifetime Use | Median Recent Use | Mean Recent Use | SD Recent Use | IQR Recent Use |
|---|---|---|---|---|---|---|---|---|---|---|---|---|
| LSD | 125 | 144.64 | 100.34 | 166.75 | 133 | 265.33 | 276.26 | 489.5 | 23.0 | 30.94 | 27.41 | 49.00 |
| Psilocybin | 18 | 28.45 | 26.75 | 48 | 126.5 | 242.60 | 260.91 | 434.75 | 26.5 | 33.59 | 28.54 | 51.25 |
| Mescaline | 382 | 385.89 | 236.08 | 371 | 278.5 | 320.86 | 254.61 | 438.75 | 34.5 | 37.82 | 26.88 | 44.50 |
| DMT | 33.5 | 37.57 | 22.93 | 38 | 260.0 | 304.72 | 258.02 | 448.00 | 36.5 | 35.92 | 26.13 | 48.00 |
| 5-MeO-DMT | 39.5 | 40.07 | 25.29 | 47 | 330.0 | 361.29 | 255.40 | 424.50 | 37.0 | 39.27 | 25.64 | 36.50 |
| Ibogaine | 37 | 39.22 | 24.3 | 41 | 345.0 | 361.32 | 245.20 | 406.50 | 29.0 | 36.61 | 23.70 | 37.00 |
| DOI | 4 | 4.45 | 2.43 | 4 | 372.0 | 390.11 | 259.80 | 447.75 | 37.5 | 42.21 | 27.67 | 51.75 |
| MDMA | 100 | 94.56 | 51.52 | 80 | 177.0 | 276.39 | 262.45 | 446.50 | 28.0 | 38.36 | 30.85 | 53.00 |
| Scopolamine | 43.5 | 42.81 | 25.37 | 42 | 396.0 | 398.50 | 259.61 | 449.00 | 35.5 | 42.61 | 29.31 | 50.00 |
| Salvia | 441.5 | 418.63 | 241.33 | 406.25 | 268.5 | 318.39 | 261.65 | 424.25 | 32.0 | 35.77 | 25.21 | 38.00 |
| Ketamine | 66 | 75.18 | 48.78 | 80 | 240.0 | 301.56 | 263.40 | 454.50 | 27.0 | 31.39 | 24.51 | 30.00 |
| PCP | 22 | 21.06 | 12.57 | 23 | 381.0 | 392.35 | 257.46 | 446.25 | 40.5 | 39.63 | 26.03 | 45.00 |

Drug assessment of classic psychedelics and substances with hallucinogenic properties including median dose (participants had to state the median dose out of all doses ever taken) and mean dose (mean dose out of all doses ever ingested). SD: Standard deviation; IQR: Interquartile range; LSD: lysergic acid diethylamide; DMT: N,N-dimethyltryptamine; 5-MeO-DMT: tryptamine; DOI: 2,5-Dimethoxy-4-iodoamphetamine; MDMA: methylenedioxymethamphetamine; PCP: phencyclidine; Lifetime use: frequency of drug use accumulated over lifetime; Recent use: frequency of use in the week prior to study participation. The dose of LSD was given in mcg; the dose of psilocybin were given in g (participants had to state weight of dried mushrooms); Doses of mescaline, DMT, 5-Me0-DMT, ibogaine, DOI, MDMA, scopolamine, salvia, ketamine, and PCP were given in mg.

**Supplemental Table S3. Descriptive Results of 10 main outcome parameters (accuracy and mean reaction time) for the five cognitive tasks completed by non-users, lifetime users, and recent users.**

| Variable | Non-user | Lifetime user | Recent user | Median Non-user | Median Lifetime user | Median Recent user | Min Non-user | Min Lifetime-user | Min Recent-user | Max Non-user | Max Lifetime-user | Max Recent user |
|---|---|---|---|---|---|---|---|---|---|---|---|---|
| Accuracy dot-probe | 144 | 181 | 434 | 13.00 (7.00 - 17.00) | 17.00 (16.00 - 17.00) | 8.00 (4.00 - 16.00) | 1 | 1 | 1 | 17 | 17 | 17 |
| Accuracy Digit span | 144 | 181 | 434 | 5.00 (3.00 - 7.00) | 7.00 (5.00 - 8.00) | 5.00 (3.00 - 7.00) | 1 | 1 | 1 | 10 | 11 | 11 |
| Accuracy N-Back | 144 | 181 | 434 | 15.00 (8.00 - 23.25) | 26.00 (18.00 - 31.00) | 9.00 (3.00 - 18.00) | 1 | 1 | 1 | 38 | 38 | 38 |
| Accuracy 2D-Rotation | 144 | 181 | 434 | 8.00 (6.00 - 10.00) | 11.00 (9.00 - 13.00) | 7.00 (4.00 - 9.00) | 1 | 3 | 1 | 16 | 16 | 16 |

| | | | | | | | | | | | | |
|---|---|---|---|---|---|---|---|---|---|---|---|---|
| Accuracy Visual Search | 144 | 181 | 434 | 10.00 (7.00 - 15.50) | 18.00 (9.00 - 20.00) | 9.00 (5.00 - 12.00) | 1 | 1 | 1 | 21 | 21 | 21 |
| Reaction Time Digit span | 144 | 181 | 434 | 4332.40 (3711.91 - 5155.62) | 3994.20 (3355.47 - 4826.86) | 4185.75 (3275.07 - 5524.29) | 1562.8 | 2237.1 | 1690 | 9869.4 | 8543.6 | 9806.8 |
| Reaction Time Dot-probe | 144 | 181 | 434 | 756.95 (496.31 - 1042.71) | 439.95 (392.93 - 519.84) | 731.43 (488.69 - 1021.12) | 202.8 | 242.7 | 200.1 | 2795.5 | 1786.8 | 3375.5 |
| Reaction Time N-Back | 144 | 181 | 434 | 855.18 (703.14 - 1049.84) | 801.61 (714.69 - 943.81) | 815.77 (610.51 - 1002.59) | 203.3 | 415.3 | 203.9 | 1473.9 | 1706.7 | 1873.4 |
| Reaction Time 2D-Rotation | 144 | 181 | 434 | 2643.53 (1703.96 - 3381.33) | 2988.33 (2481.30 - 3671.50) | 2022.96 (979.08 - 2935.38) | 215.55 | 655.775 | 213.65 | 5735.182 | 6119.817 | 8406 |
| Reaction Time Visual Search | 144 | 181 | 434 | 1014.16 (671.30 - 1502.79) | 1021.83 (781.11 - 1246.58) | 1017.70 (671.41 - 1516.11) | 206.5667 | 323.5 | 203.5 | 3592.389 | 4989.4 | 4104.833 |

**Supplemental Table S4. Recent Drug Use x Cognitive Tasks**

| Task | Variable | Estimate | *P* Value | Test Statistic |
|---|---|---|---|---|
| Dot-probe | Reaction Time | 0.248 | < 0.001 | 5.435 |
| Dot-probe | Accuracy | -0.308 | < 0.001 | -6.874 |
| 2D-Rotation | Reaction Time | -0.24 | 0.00000024 | -5.246 |
| 2D-Rotation | Accuracy | -0.331 | < 0.001 | -7.452 |
| Visual Search | Reaction Time | -0.019 | 0.694 | -0.393 |
| Visual Search | Accuracy | -0.173 | 0.000208 | -3.739 |
| N-Back | Reaction Time | 0.014 | 0.759 | 0.307 |
| N-Back | Accuracy | -0.305 | < 0.001 | -6.814 |
| Digit Span | Reaction Time | 0 | 0.994 | 0.007 |
| Digit Span | Accuracy | -0.119 | 0.0114 | -2.54 |

**Lifetime Drug Use x Cognitive Tasks**

| Task | Variable | Estimate | *P* Value | Test Statistic |
|---|---|---|---|---|
| Dot-probe | Reaction Time | 0.061 | 0.197 | 1.291 |
| Dot-probe | Accuracy | -0.155 | 0.000932 | -3.332 |
| Rotation | Reaction Time | -0.187 | 0.0000598 | -4.052 |
| Rotation | Accuracy | -0.166 | 0.00038 | -3.581 |
| Visual Search | Reaction Time | -0.005 | 0.91 | -0.113 |
| Visual Search | Accuracy | -0.119 | 0.011 | -2.554 |
| N-Back | Reaction Time | -0.026 | 0.582 | -0.551 |

| | | | | |
|---|---|---|---|---|
| N-Back | Accuracy | -0.223 | 0.0000017 | -4.85 |
| Digit Span | Reaction Time | 0.002 | 0.967 | 0.041 |
| Digit Span | Accuracy | -0.013 | 0.777 | -0.283 |

Above: Partial Spearman Correlations of consumed psychedelics and substances with hallucinogenic properties in the week prior to task performance (recent use) and outcomes (accuracy and mean reaction time) of five cognitive tasks (dot-probe task, 2D mental rotation task, visual search task, n-back task, digit span text entry task). Coefficients between recent psychedelic use and cognitive performance, controlling for age at first use, sex, education level, age, use of non psychedelic drugs, and psychiatric diagnosis. Positive estimates indicate associations with slower reaction times or worse performance, while negative estimates indicate better accuracy or faster responses. Significant correlations ($p < 0.05$) suggest potential negative cognitive effects for accuracy across all five tasks. Below: Same as above but for participants who stated taking any psychedelic drug or substance with hallucinogenic properties in their lifetime (lifetime users). Significant correlations were found for negative effects on accuracy for the dot-probe, 2D mental rotation, visual search, and n-back tasks. Correlations for mean reaction time were mixed.

| **LSD** | | | | **Psilocybin** | | | |
|---|---|---|---|---|---|---|---|
| **Task** | **Variable** | **Spearman** | ***P* value** | **Task** | **Variable** | **Spearman** | ***P* value** |
| Dot-probe | Accuracy | -0.193 | 0.00000387 | Dot-probe | Accuracy | -0.541 | < 0.001 |
| Dot-probe | Reaction Time | 0.0933 | 0.0270 | Dot-probe | Reaction Time | 0.319 | < 0.001 |
| 2D-Rotation | Accuracy | -0.185 | 0.00000375 | 2D-Rotation | Accuracy | -0.500 | < 0.001 |
| 2D-Rotation | Reaction Time | 0.161 | 0.0000584 | 2D-Rotation | Reaction Time | -0.398 | < 0.001 |
| Visual Search | Accuracy | -0.144 | 0.000481 | Visual Search | Accuracy | -0.433 | < 0.001 |
| Visual Search | Reaction Time | 0.00304 | 0.941 | Visual Search | Reaction Time | -0.0314 | 0.448 |
| N-Back | Accuracy | -0.233 | 0.0000000199 | N-Back | Accuracy | -0.539 | < 0.001 |
| N-Back | Reaction Time | -0.00586 | 0.889 | N-Back | Reaction Time | -0.00789 | 0.851 |
| Digit Span | Accuracy | -0.116 | 0.00694 | Digit Span | Accuracy | -0.303 | < 0.001 |
| Digit Span | Reaction Time | 0.0245 | 0.569 | Digit Span | Reaction Time | 0.0701 | 0.103 |

| **Alcohol** | | | | **THC** | | | |
|---|---|---|---|---|---|---|---|
| **Task** | **Variable** | **Spearman** | ***P* value** | **Task** | **Variable** | **Spearman** | ***P* value** |
| Dot-probe | Accuracy | -0.517 | < 0.001 | Dot-probe | Accuracy | -0.427 | < 0.001 |
| Dot-probe | Reaction Time | 0.340 | < 0.001 | Dot-probe | Reaction Time | 0.219 | < 0.001 |
| 2D-Rotation | Accuracy | -0.470 | < 0.001 | 2D-Rotation | Accuracy | -0.376 | < 0.001 |
| 2D-Rotation | Reaction Time | -0.321 | < 0.001 | 2D-Rotation | Reaction Time | -0.306 | < 0.001 |
| Visual Search | Accuracy | -0.379 | < 0.001 | Visual Search | Accuracy | -0.332 | < 0.001 |

| | | | | | | | |
|---|---|---|---|---|---|---|---|
| Visual Search | Reaction Time | -0.0320 | 0.389 | Visual Search | Reaction Time | -0.0408 | 0.272 |
| N-Back | Accuracy | -0.455 | < 0.001 | N-Back | Accuracy | -0.395 | < 0.001 |
| N-Back | Reaction Time | 0.0122 | 0.746 | N-Back | Reaction Time | -0.0708 | 0.0609 |
| Digit Span | Accuracy | -0.306 | < 0.001 | Digit Span | Accuracy | -0.233 | < 0.001 |
| Digit Span | Reaction Time | 0.0609 | 0.117 | Digit Span | Reaction Time | 0.0272 | 0.484 |

**Supplemental Table S5: Corrected Spearman correlations for dose-dependent accuracy and reaction time outcomes for LSD, psilocybin, alcohol and THC**

Correlations were calculated separately for LSD, psilocybin, alcohol, and THC, and for each of five cognitive tasks (dot-probe, 2D mental rotation, visual search, n-back, and digit span text entry), assessing accuracy and reaction time. Spearman's rho values and corresponding Bonferroni-corrected *p*-values are reported. Dose-dependent effects were observed for LSD, psilocybin, alcohol, and THC for accuracy across all five cognitive tasks, with higher median doses correlated with lower accuracy; outcomes for reaction time varied.

| Questionnaire | Non-user | Lifetimeuser | Recentuser | Median Non-user | Median Lifetime user | Median recent user | Min Non-user | Min Lifetimeuser | Min Recentuser | Max Non-user | Max Lifetimeuser | Max Recentuser |
|---|---|---|---|---|---|---|---|---|---|---|---|---|
| DES-B | 144 | 181 | 434 | 20.00 (13.00 - 28.25) | 13.00 (12.00 - 18.00) | 29.00 (25.00 - 33.00) | 8 | 8 | 8 | 36 | 34 | 40 |
| HPPD | 144 | 181 | 434 | 22.00 (15.00 - 32.00) | 15.00 (13.00 - 18.00) | 36.00 (30.00 - 40.00) | 12 | 12 | 12 | 44 | 38 | 48 |
| PHQ-9 | 144 | 181 | 434 | 20.00 (13.00 - 24.25) | 14.00 (11.00 - 19.00) | 25.00 (22.00 - 29.00) | 9 | 9 | 9 | 34 | 32 | 35 |
| WHO-QOL-Overall | 144 | 181 | 434 | 16.00 (14.00 - 18.00) | 16.00 (12.00 - 18.00) | 16.00 (14.00 - 18.00) | 6 | 6 | 4 | 20 | 20 | 20 |

| | | | | | | | | | | | |
|---|---|---|---|---|---|---|---|---|---|---|---|
| Environmental Domain | 144 | 181 | 434 | 15.75 (14.00 - 17.00) | 15.50 (14.00 - 17.00) | 15.50 (14.00 - 17.00) | 10 | 7,5 | 7 | 20 | 20 | 20 |
| Physical Domain | 144 | 181 | 434 | 14.29 (13.14 - 15.43) | 14.86 (13.14 - 17.14) | 13.71 (13.14 - 14.86) | 6.86 | 8.57 | 8.57 | 20 | 20 | 20 |
| Psychological Domain | 144 | 181 | 434 | 14.33 (13.17 - 16.17) | 14.00 (12.00 - 15.33) | 17.67 (13.33 - 15.33) | 5.33 | 5.33 | 6.67 | 20 | 20 | 19.33 |
| Social Domain | 144 | 181 | 434 | 16.00 (13.33 - 17.33) | 13.33 (10.67 - 16.00) | 16.00 (13.33 - 17.33) | 4 | 4 | 4 | 20 | 20 | 20 |

**Supplemental Table S6. Descriptive Results for four mental health questionnaires grouped by non-user, lifetime user, and recent user**

DES-B: Brief Dissociative Experiences Scale (highest possible score: 40; higher score indicates more intense/frequent dissociative symptoms; HPPD: Hallucinogen Persistent Perception Disorder (highest possible score: 48, higher score indicates more intense/frequent HPPD symptoms); PHQ-9: nine-item Patient Health Questionnaire (highest possible score: 36, higher score indicates more severe depressive symptoms); WHO-

QOL: World Health Organization - Quality of Life questionnaire. The WHO-QOL questionnaire is divided into five parts: overall quality of life and physical, psychological, social, and environmental domains (highest score for each domain: 20, higher scores indicate higher quality of life in each domain).

**Supplemental Table S7: Explorative Analysis: Kruskal-Wallis test for five cognitive tasks and Mental Health Questionnaires excluding "currently-using" subjects of the lifetime user group**

| Dependent Variable | Test Statistic | *p*-Value |
|---|---|---|
| Reaction Time Dotprobe | 107,738 | < 0.001 |
| Accuracy Dotprobe | 169,038 | < 0.001 |
| Reaction Time 2D-Rotation | 109,879 | < 0.001 |
| Accuracy 2D-Rotation | 190,147 | < 0.001 |
| Reaction Time Visual Search | 1,379 | 0.5018 |
| Accuracy Visual Search | 84,219 | < 0.001 |
| Reaction Time N-back | 3,321 | 0.19 |
| Accuracy N-back | 145,591 | < 0.001 |
| Reaction Time Digit span | 4,516 | 0.1046 |
| Accuracy Digit span | 49,824 | < 0.001 |
| PHQ-9 | 200,642 | < 0.001 |
| HPPD | 258,724 | < 0.001 |
| Physical Domain | 57,082 | < 0.001 |
| Psychological Domain | 12,116 | 0.0023 |
| Social Domain | 43,683 | < 0.001 |
| Environmental Domain | 0,248 | 0.8836 |
| WHO-QOL-Overall | 11,53 | 0.0031 |
| DES-B | 240,302 | < 0.001 |

Explorative analysis of Kruskal-Wallis test results comparing cognitive and mental health outcomes between three groups based on psychedelic use (use within last week, lifetime use, and never used). Participants stating lifetime use and that they were under the influence of a psychedelic drug during study participation, but did not state psychedelic drug use in the week prior to study participation (n=23) and were hence part of the lifetime user group were excluded from analysis. Reported are the test statistics ($\chi^2$) and corresponding p-values. Significant results ($p < .05$) indicate group differences for the respective outcome variables.

**Supplemental Table S8: Spearman Correlation of Lifetime Drug use (excluding "currently-using" users of the lifetime group) x cognitive tasks**

| Task | Variable | Estimate | *P*-Value | Statistic |
|---|---|---|---|---|
| Dotprobe | Mean Reaction Time | 0,074 | 0,125 | 1,536 |
| Dotprobe | Correct Responses | -0,245 | < 0.001 | -5,261 |
| Rotation | Mean Reaction Time | -0,239 | < 0.001 | -5,122 |
| Rotation | Correct Responses | -0,243 | < 0.001 | -5,215 |
| Visual Search | Mean Reaction Time | -0,03 | 0,539 | -0,615 |
| Visual Search | Correct Responses | -0,163 | < 0.001 | -3,423 |
| N-back | Mean Reaction Time | -0,055 | 0,251 | -1,149 |
| N-back | Correct Responses | -0,277 | < 0.001 | -6,001 |
| Digit Span | Mean Reaction Time | 0,003 | 0,946 | 0,068 |
| Digit Span | Correct Responses | -0,06 | 0,214 | -1,245 |

Explorative analysis: Partial Spearman correlation analyses between lifetime use of hallucinogens and cognitive outcome measures, controlling for covariates (age at first use, sex, education level, current age, use of non-hallucinogens, and psychiatric diagnosis). Participants stating lifetime use and that they were under the influence of a psychedelic drug during study participation, but did not state psychedelic drug use in the week prior to study participation (n=23) and were hence part of the lifetime user group were excluded from analysis. Reported are correlation estimates, test statistics, p-values, and sample sizes (N).

Significant negative correlations indicate worse cognitive performance with increasing lifetime hallucinogen use.

**Supplemental Figure S1. Additional Variables of Psychedelic Use.**

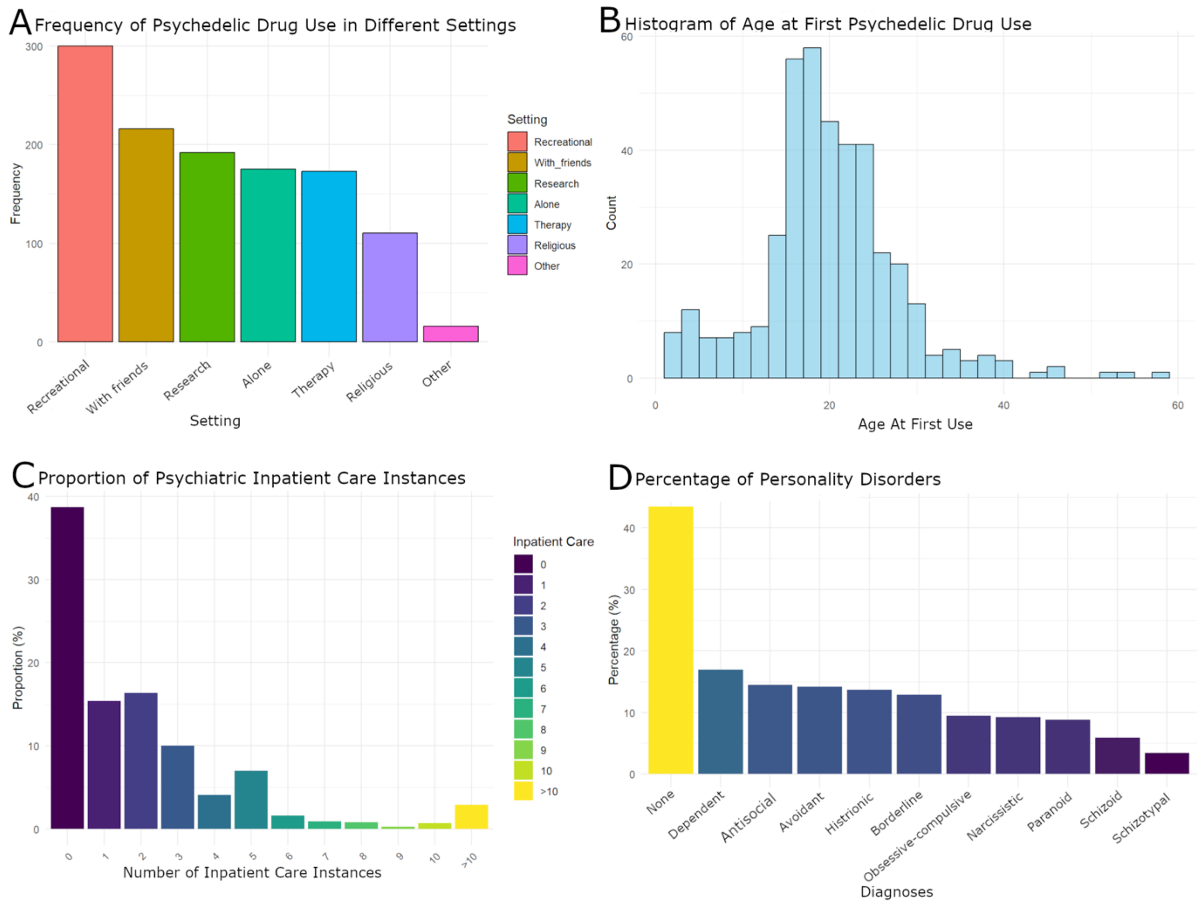


A: The frequency of different settings in which participants reported psychedelic use. B: Age at first psychedelic drug use. C: Number of psychiatric inpatient care instances. Most participants had never been hospitalized. D: Proportion of personality disorder diagnoses among participants.

**Supplemental Figure S2. Partial Spearman Correlation between drug use and cognitive performance**

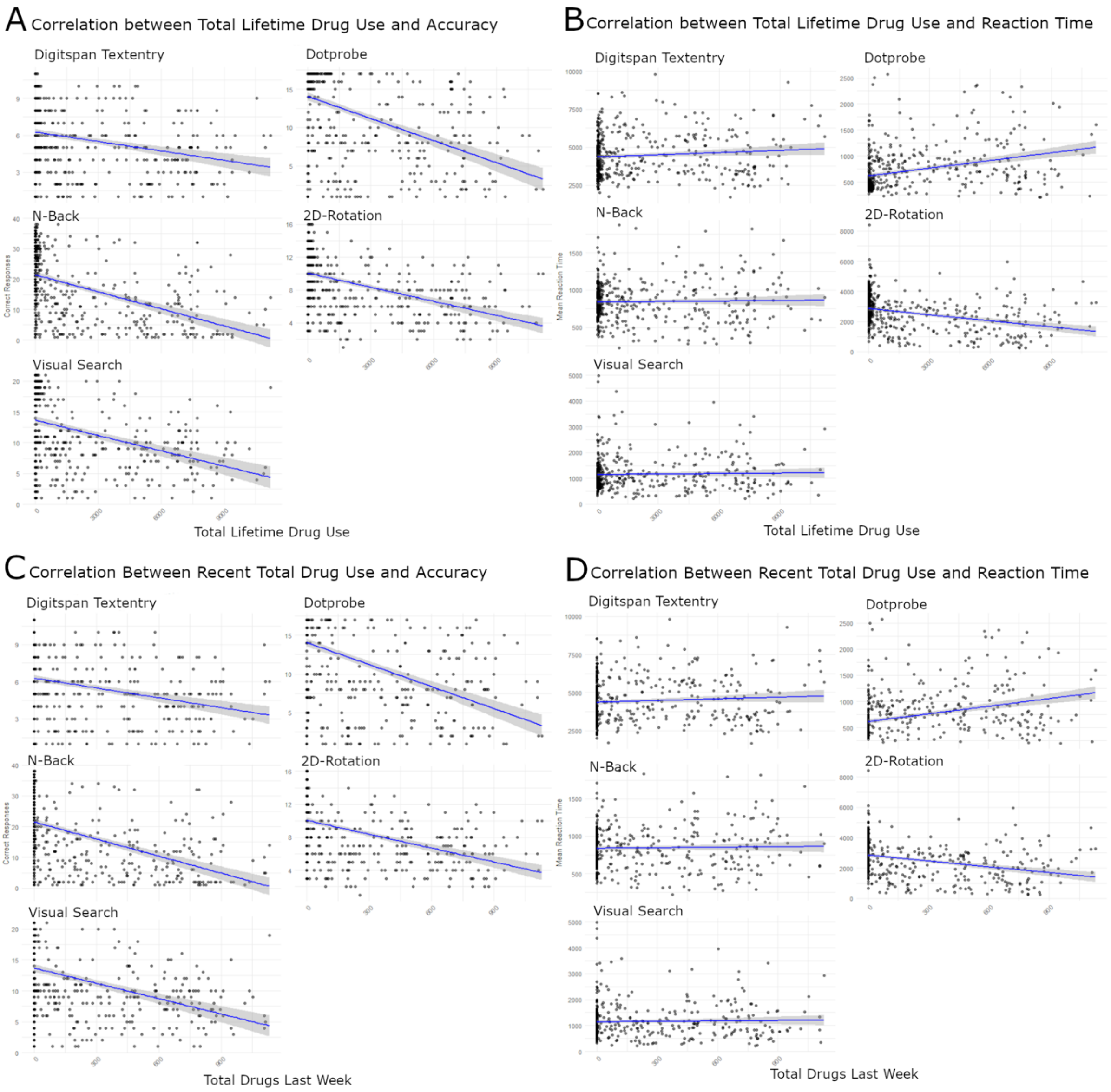


**A:** Correlation between total lifetime drug use and task accuracy across five cognitive tasks: digit span text entry, dot-probe, n-Back, 2D-rotation, and visual search. **B:** Correlation between total lifetime drug use and mean reaction time across the same tasks. **C:** Correlation between recent (within week prior to study participation) total drug use and task accuracy. **D:** Correlation between recent total drug use and mean reaction time. All plots depict partial

Spearman correlation coefficients controlling for age, sex, and education. Blue regression lines represent the correlation trend with 95% confidence intervals shaded.
